# The impact of evolving maternal antiretroviral therapy guidelines on vertical transmission of HIV in the Western Cape, South Africa

**DOI:** 10.1101/2025.07.06.25330971

**Authors:** Nisha Jacob, Emma Kalk, Alexa Heekes, Florence Phelanyane, Kim Anderson, Mary-Ann Davies, Brian Rice, Andrew Boulle

**Author notes:** **Corresponding author:** Nisha Jacob School of Public Health, Faculty of Health Sciences University of Cape Town Anzio Road, Observatory 7925.

## Abstract

**Introduction:** Despite universal HIV test-and-treat policy in South Africa, vertical transmission continues to occur. We evaluated the temporal effectiveness of vertical transmission prevention (VTP) over three maternal antiretroviral therapy (ART) policy periods (three-drug ART accessible to those with CD4 counts <200 cells/µl (January 2010 – March 2010); <350 cells/µl (April 2010 – April 2013); and lifelong ART regardless of CD4 count (May 2013 – December 2020)) in Western Cape, South Africa using public sector routine individuated data.

**Methods:** We conducted a retrospective cohort study with child HIV infection as the primary outcome. The cohort of 842 641 pregnancies from 2010 – 2020, inclusive of child HIV exposure, maternal ART, and child outcomes, was enumerated using administrative, laboratory and pharmacy data. Multivariable logistic regression was used to explore associations with vertical transmission.

**Results:** The proportion of pregnant women living with HIV (WLWH) initiating ART prior to pregnancy increased from 20.9% in 2010 to 71.1% in 2020. Of all pregnancies 17.1% (143 987/842 641) were HIV exposed and 16.3% (137 572/842 641) had a record of a child HIV outcome, of whom 3966 (2.9%) were HIV positive by 24 months. Among children with known maternal HIV exposure (143 987), 32.0% had unknown HIV status and 2.1% were diagnosed with HIV by study closure. In 2020, HIV status was ascertained in 87.2% (16 908/19 382) of children exposed to HIV. Children born in policy period 3 were less likely to have HIV than children born in policy period 2 (aOR 0.66; 95% CI 0.60 – 0.72), mediated through expanded ART access. Between 2017 and 2020, 16.5% of child HIV diagnoses were from pregnancies without maternal HIV exposure records. Young maternal age, no antenatal ART, previous tuberculosis and no records of antenatal visits were associated with vertical transmission in all periods.

**Conclusions:** Using routine data, we report an increase in WLWH initiating ART prior to pregnancy, and a decline in vertical transmission of HIV over three policy periods. Although HIV diagnosis amongst children exposed to HIV has reduced over time, the proportion of infections in children without established exposure emphasises the need to optimise VTP opportunities.

## Introduction

Vertical transmission (VT) of HIV from pregnant women living with HIV (WLWH) to their children is a preventable adverse outcome of HIV in pregnancy and carries considerable risk of paediatric morbidity and mortality[1,2]. The WHO Initiative for elimination of vertical transmission of HIV highlights the importance of early antenatal care for all WLWH, early HIV diagnosis and antiretroviral therapy (ART) initiation[3,4]. Despite the implementation of lifelong ART for all pregnant WLWH (option B+) and later Universal Test and Treat in South Africa, VT continues to occur during pregnancy, delivery and during the post-partum period, if breastfeeding[5,6].

As ART access in South Africa progressed from 1999, several large studies were conducted using routine data to highlight the impact of ART and vertical transmission prevention (VTP) programmes on maternal and child outcomes and the need for scale up and revision of guidelines, both internationally and locally[7–13]. In the current universal ART era, few studies have used routine data to evaluate vertical transmission throughout pregnancy and post-partum. This may be due to challenges of accessing complete routine programmatic data. More recent studies in the Western Cape province, South Africa have been based on case histories from single facilities or limited to a few selected facilities[1,14–16]. Although these studies provide valuable insights, results have limited external validity for population-wide inference and action.

The Western Cape Provincial Government in South Africa has developed a Provincial Health Data Centre (PHDC) in which all public-sector electronic individual-level routine data are consolidated on a single platform, enabling the description of temporal changes in population-wide outcomes and risk factors for VT along the VTP cascade (an evolving set of sequential steps in the health services pathway to prevent VT[17]. Using routine province-wide individuated data, we evaluated the temporal effectiveness of VTP over different maternal ART and infant polymerase chain reaction (PCR) testing policy periods since 2010, and associations with transmission.

## Methods

### Study Design and Population

The study was set in the Western Cape province, South Africa, which is comprised of one urban district, the Cape Metropolitan, and five less urbanised districts, Overberg, Garden Route, Central Karoo, West Coast and Cape Winelands. We conducted a retrospective cohort study including all pregnancies of women attending public health facilities within the Western Cape from January 2010 to December 2020 with child HIV infection as the primary outcome.

### Data Sources and Processing

The study cohort was enumerated using the PHDC Maternity Cascade which links electronic records of all patients with electronic administrative or laboratory evidence indicative of pregnancy. De-identified data were accessed on 25/05/2022. Pregnancies were identified by a rhesus antibody test (conducted routinely at first antenatal visit), a pregnancy test, birth register record, antenatal visit or maternal discharge summary. Only pregnancies with live birth outcomes were included. Mother-child pairs were electronically linked by the PHDC.

The PHDC HIV care cascade was linked to the maternity care cascade to determine maternal and child HIV status and associated HIV interventions (Figure 1).

**Figure 1:**
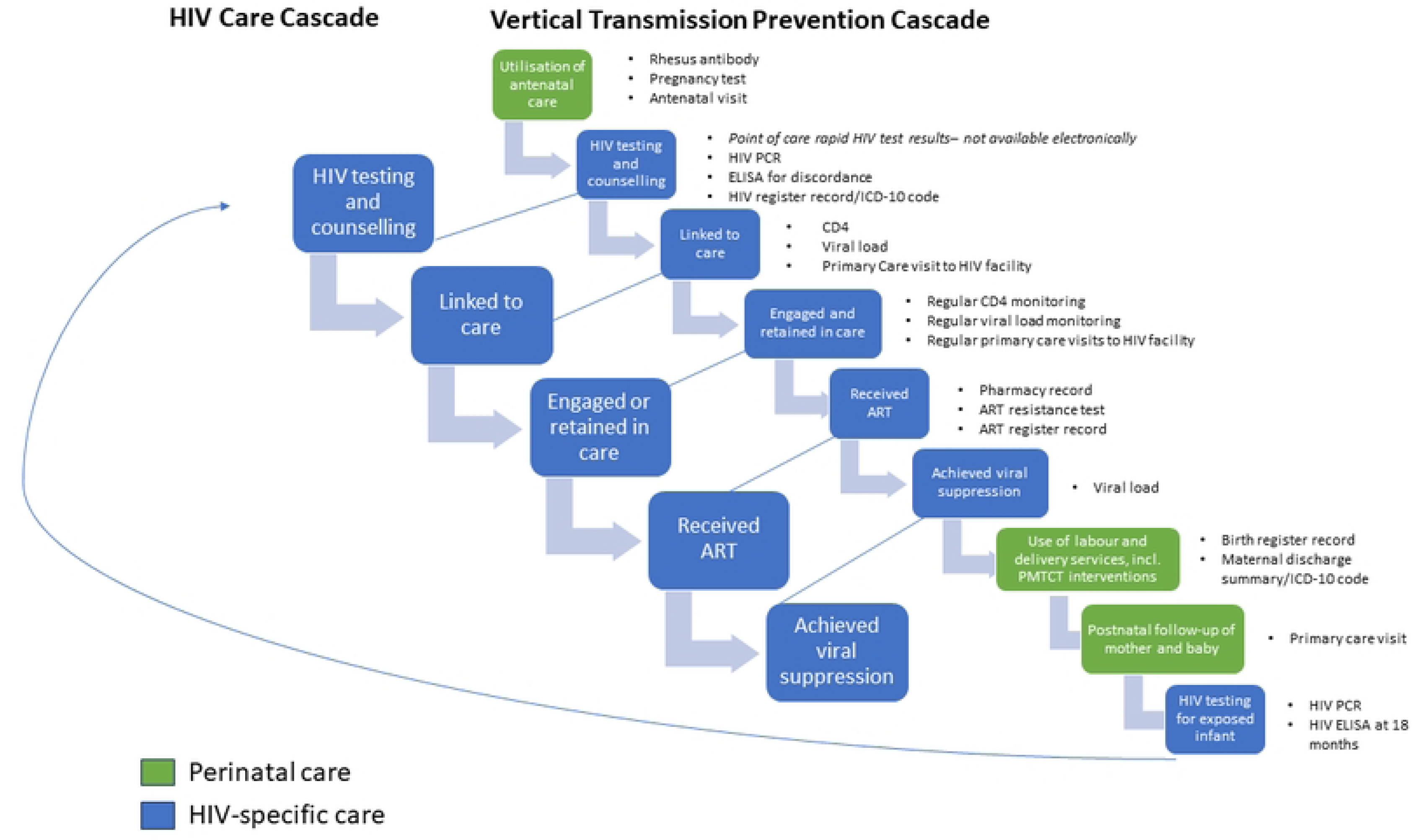
PHDC HIV and VTP Cascade data points.

The first step of the HIV care cascade, diagnosis of HIV, is typically instigated following positive point-of-care tests for adults. By protocol, all pregnant women not known to have HIV, are offered rapid point-of-care HIV testing on their first antenatal presentation to a health facility[18]. Since 2013, repeat HIV testing with point-of-care HIV rapid assays should be offered to women with negative/unknown HIV status, in the second and third trimesters, in labour or immediately after delivery, at 6 weeks after delivery and every 3 months while breastfeeding[18,19]. These results are not routinely digitised and are therefore unavailable to the PHDC. Maternal HIV positive PHDC status is ascertained if laboratory testing shows a detectable viral load, positive HIV ELISA test, a test for ART resistance or evidence of initiating ART in either pharmacy or administrative information systems, HIV register record or HIV-related International Classification of Diseases (ICD) Tenth Revision code.

Over time, various policy changes have been implemented which impacted routine data available to the PHDC. For example, changes in CD4 and viral load (VL) monitoring in pregnant women took place in 2013 with VL testing becoming the preferred tool for treatment monitoring and follow-up CD4 testing being no longer conducted routinely[18–20]. Notable policy changes to ART initiation for WLWH and HIV testing of children HIV exposed are shown in Figure 2 [21,22].

**Figure 2:**
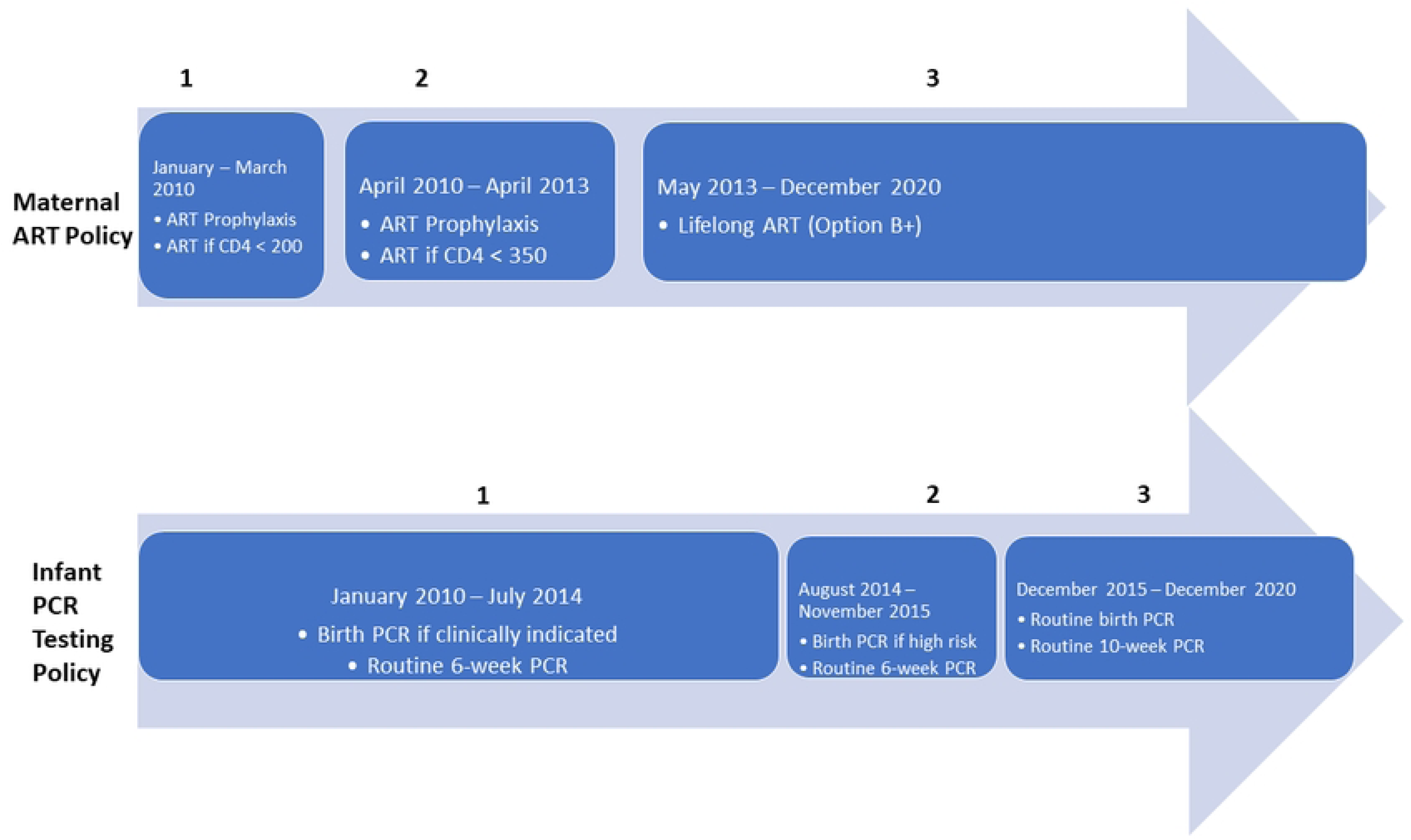
Changes in Maternal antiretroviral therapy (ART) and Infant HIV PCR testing policies, Western Cape, South Africa 2010 – 2020.

Infants exposed to HIV are tested for HIV using an HIV PCR test (Figure 2). From 2015 onwards, routine infant PCR tests were conducted in all infants exposed to HIV within 48 hours of birth, at 10 weeks and 18 weeks (only if breastfeeding and received Nevirapine for 12 weeks), followed by a rapid HIV test at 9 months with confirmatory PCR test if positive. From 2020, routine tests at 18 weeks were replaced by a PCR test at 6 months[18,19]. These results are digitised in the National Health Laboratory System (NHLS) and integrated by the PHDC, providing reliable routine evidence of HIV status of infants in addition to other HIV-specific routine data such as viral load testing and ART.

HIV exposure was defined as a child whose mother had evidence of HIV antenatally or up to two years from delivery, as breastfeeding is more likely to be discontinued by 24 months[23]. Children whose mothers had no evidence of HIV up to two years from delivery were considered to have unknown exposure. For multiple pregnancies (e.g. twins and triplets), only the first child’s HIV outcome was assigned to the pregnancy. Vertical transmission diagnosis was defined as “early” if a child’s HIV diagnosis was recorded at or before seven weeks of age, and “late” if diagnosis was after seven weeks of age.

### Analysis

Data were analysed using Stata 17 (Stata Corporation, College Station, Texas, USA). Measures of central tendency and dispersion were used to describe continuous variables, depending on distribution. Categorical variables were described using proportions and 95% confidence intervals. Viral load and CD4 test results within one year before and after the estimated pregnancy start date were categorised according to Western Cape protocol-related result thresholds[18,19]. Viral load status was considered viraemic or suppressed if the last available VL, within 1 year of estimated pregnancy start date, was greater (inclusive) or less than 1000 copies/ml, respectively. Data were further analysed according to maternal ART policy period as described in Figure 1 [22,24], with 2019 descriptive characteristics also presented as the most recent pre-COVID-19 patient profile within the dataset. Vertical transmission characteristics were analysed for the full cohort period and a shorter three-year period (2017 – 2020) to reflect more recent characteristics under updated policy guidelines.

Routine data on antenatal care, maternal and child HIV care, ART and other descriptive characteristics were included for multivariable analyses of risk factors for vertical transmission. A directed acyclic graph was drawn using DAGitty v3.1 to explore causal relationships and select variables available from routine data for model building, addressing collider, collinear and confounder effects. Electronic evidence of prior pregnancy was excluded from model building due to incomplete electronic systems in earlier years precluding ascertainment of pregnancy. Four logistic regression models were built to demonstrate the impact of selected variables. Model 1 included maternal ART policy period without adjustment for maternal ART status and associated laboratory outcomes. Model 2 included both maternal ART policy period and maternal ART status, to explore potential mediation by ART status. Models 3 and 4 were restricted to policy period 3 to demonstrate the effect of VL and CD4 independently due to collinearity with policy period. Model 3 included VL status combined with ART status into a composite variable. Model 4 included combined VL and ART status as well as CD4 status. Unadjusted and adjusted odd ratios are reported. For variables with missing data, a missing data category was included. This was preferred to imputation methods as these are not data missing at random, and the fact of missingness is of interest.

### Ethical considerations

The study was approved by the University of Cape Town Human Research Ethics Committee (HREC 083/2021) and the Western Cape Provincial Health Research Committee. The research followed the ethical standards outlined in the Declaration of Helsinki. Data from the PHDC are unconsented, de-identified data housed by the Western Cape Department of Health and Wellness. Data were fully de-identified before release for the study according to the Western Cape Department of Health Data Access Policy Guidelines.

## Results

Between 2010 and 2020, 842 641 pregnancies were included in the study (Table 1). Most pregnancies (70.5%) were recorded in the Cape Metro district, with the proportion differing by policy period (Table 1). The proportion of pregnancies among women diagnosed with HIV prior to or during pregnancy was higher in maternal ART period 3 (17.2%) than in periods 1 (9.2%) or 2 (10.9%).

**Table 1:** Descriptive characteristics of pregnancy cohort by Maternal ART Policy Period.

| Variables | Maternal ART Policy Period 1<br>Jan – Mar 2010<br>(n=9 946)<br>1.2% |  | Maternal ART Policy Period 2<br>Apr 2010 – Apr 2013<br>(n= 140 710)<br>16.7% |  | Maternal ART Policy Period 3<br>May 2013 - Dec 2020<br>(n=691 985)<br>82.1% |  | 2019<br>(n = 99 055) |  | Total<br>2010 - 2020<br>(N= 842 641) |  |
| --- | --- | --- | --- | --- | --- | --- | --- | --- | --- | --- |
|  | n | % | n | % | n | % | n | % | n | % |
| **District** |  |  |  |  |  |  |  |  |  |  |
| Cape Winelands | 999 | 10.0 | 16412 | 11.7 | 95132 | 13.8 | 12858 | 13.0 | 112543 | 13.4 |
| Central Karoo | 10 | 0.1 | 52 | 0.0 | 6246 | 0.9 | 1234 | 1.3 | 6308 | 0.8 |
| City of Cape Town Metro | 8626 | 86.7 | 114552 | 81.4 | 471183 | 68.1 | 64891 | 65.5 | 594361 | 70.5 |
| Garden Route | 284 | 2.9 | 9126 | 6.5 | 61511 | 8.9 | 9242 | 9.3 | 70921 | 8.4 |
| Overberg | 10 | 0.1 | 191 | 0.1 | 23127 | 3.3 | 4220 | 4.3 | 23328 | 2.8 |
| West Coast | 3 | 0.0 | 90 | 0.06 | 29770 | 4.3 | 5476 | 5.5 | 29863 | 3.5 |
| Unknown | 14 | 0.1 | 287 | 0.2 | 5016 | 0.7 | 1134 | 1.1 | 5317 | 0.6 |
| **Maternal age (median, IQR in years)** | 26.3 (22.1 – 31.4) |  | 26.4 (22.1 – 31.2) |  | 26.8 (22.4 – 31.7) |  | 27.0 (22.7 – 32.1) |  | 26.7 (22.4 – 31.6) |  |
| **Electronic evidence of prior pregnancy** |  |  |  |  |  |  |  |  |  |  |
| 0 | 7929 | 79.7 | 103011 | 73.2 | 405737 | 58.6 | 51827 | 52.3 | 516677 | 61.3 |
| 1 | 1733 | 17.4 | 31043 | 22.1 | 192283 | 27.8 | 29440 | 29.7 | 225059 | 26.7 |
| 2 | 248 | 2.5 | 5761 | 4.1 | 68704 | 9.9 | 12268 | 12.4 | 74713 | 8.9 |
| 3 | 33 | 0.3 | 773 | 0.6 | 19244 | 2.8 | 4021 | 4.1 | 20050 | 2.4 |
| ≥4 | 3 | 0.03 | 120 | 0.1 | 6015 | 0.9 | 1499 | 1.5 | 6138 | 0.7 |
| **Maternal HIV status in index pregnancy†** |  |  |  |  |  |  |  |  |  |  |
| Evidence of HIV - status | 996 | 10.0 | 13777 | 9.8 | 58119 | 8.4 | 7183 | 7.3 | 72892 | 8.7 |
| Evidence of HIV + status | 2056 | 20.7 | 29957 | 21.3 | 137153 | 19.8 | 18968 | 19.2 | 169166 | 20.1 |
| No HIV evidence | 6894 | 69.3 | 96976 | 68.9 | 496713 | 71.8 | 72904 | 73.6 | 600583 | 71.2 |
| **Maternal HIV timing in pregnancy period** |  |  |  |  |  |  |  |  |  |  |
| HIV + before or during pregnancy | 911 | 9.2 | 15370 | 10.9 | 118663 | 17.2 | 17912 | 18.1 | 134944 | 16.0 |
| HIV + within 2 years of delivery | 191 | 1.9 | 2582 | 1.8 | 6270 | 0.9 | 708 | 0.7 | 9043 | 1.1 |
| HIV + 2 years after delivery | 954 | 9.6 | 12005 | 8.5 | 12220 | 1.8 | 348 | 0.4 | 25179 | 3.0 |
| HIV - or unknown | 7890 | 79.3 | 110753 | 78.7 | 554832 | 80.2 | 80087 | 80.9 | 673475 | 79.9 |
| **Previous TB** | 1026 | 10.3 | 13317 | 9.5 | 54904 | 7.9 | 6943 | 7.0 | 69247 | 8.2 |
| **No recorded antenatal visits** | 3401 | 34.2 | 51952 | 36.9 | 183207 | 26.5 | 22347 | 22.6 | 238560 | 28.3 |
| **Maternal ART (n=169 166)** | **2056** |  | **29957** |  | **137153** |  | **18 968** |  | **169166** |  |
| ART started before pregnancy | 171 | 8.3 | 4596 | 15.3 | 65613 | 47.8 | 12121 | 63.9 | 70380 | 41.6 |
| ART started during pregnancy | 281 | 13.7 | 6308 | 21.1 | 44786 | 32.7 | 4836 | 25.5 | 51375 | 30.4 |
| ART started postnatally HIV + during pregnancy | 202 | 9.8 | 1722 | 5.8 | 1842 | 1.3 | 174 | 0.9 | 3766 | 2.2 |
| ART started postnatally | 1049 | 51.0 | 13785 | 46.0 | 16004 | 11.7 | 824 | 4.3 | 30838 | 18.2 |
| No ART | 353 | 17.2 | 3546 | 11.8 | 8908 | 6.5 | 1013 | 5.3 | 12807 | 7.6 |
| **Antenatal VL (copies/ml) status among those with HIV before/during pregnancy (n=134 944)** | **911** |  | **15370** |  | **118663** |  | **17 912** |  | **134944** |  |
| VL suppressed (<1000) | 190 | 20.9 | 6205 | 40.4 | 82637 | 69.6 | 12 975 | 72.4 | 89032 | 66.0 |
| Viraemic (≥1000) | 49 | 5.4 | 1092 | 7.1 | 18002 | 15.2 | 2 624 | 14.7 | 19143 | 14.2 |
| VL not available | 672 | 73.8 | 8073 | 52.5 | 18024 | 15.2 | 2 313 | 12.9 | 26769 | 19.8 |
| **Viral load status and Antenatal ART status amongst those with HIV before/during pregnancy (n=134 944)** | **911** |  | **15370** |  | **118663** |  | **17 912** |  | **134944** |  |
| Suppressed, ART before/during pregnancy | 181 | 19.9 | 5930 | 38.6 | 79149 | 66.7 | 12497 | 69.8 | 85260 | 63.2 |
| Viraemic, ART before/during pregnancy | 40 | 4.4 | 933 | 6.1 | 16700 | 14.1 | 2469 | 13.8 | 17673 | 13.1 |
| VL unavailable, ART before/during pregnancy | 231 | 25.4 | 4041 | 26.3 | 14550 | 12.3 | 1991 | 11.1 | 18822 | 14.0 |
| Suppressed, no antenatal ART record | 9 | 1.0 | 275 | 1.8 | 3488 | 2.9 | 478 | 2.7 | 3772 | 2.8 |
| Viraemic, no antenatal ART record | 9 | 1.0 | 159 | 1.0 | 1302 | 1.1 | 155 | 0.9 | 1470 | 1.1 |
| VL unavailable, no antenatal ART | 441 | 48.1 | 4032 | 26.2 | 3474 | 2.9 | 322 | 1.8 | 7947 | 6.0 |
| **Last CD4 (cells/μl) within 1 year before estimated pregnancy start among those with antenatal initiation of ART (n= 51375)** |  |  |  |  |  |  |  |  |  |  |
|  | **281** |  | **6308** |  | **44786** |  | **4836** |  | **51375** |  |
| Median (IQR) | 301 (233.9 – 342.9) |  | 336.9 (254.0 – 430.9) |  | 505 (366.0 – 671.0) |  | 455 (305.5 – 640.5) |  | 470 (332.1 – 640.0) |  |
| CD4 <200 | 6 | 2.1 | 145 | 2.3 | 381 | 0.9 | 36 | 0.7 | 532 | 1.0 |
| CD4 200 – 350 | 25 | 8.9 | 468 | 7.4 | 1006 | 2.3 | 65 | 1.3 | 1499 | 2.9 |
| CD4 >350 | 9 | 3.2 | 521 | 8.3 | 4785 | 10.7 | 211 | 4.4 | 5315 | 10.4 |
| CD4 result not available | 241 | 85.8 | 5174 | 82.0 | 38614 | 86.2 | 4524 | 93.6 | 44029 | 85.7 |
| **Last CD4 (cells/μl) among those with a CD4 test within 2 weeks of antenatal ART initiation (n=31 813)** |  |  |  |  |  |  |  |  |  |  |
|  | **24** |  | **1069** |  | **30720** |  | **3557** |  | **31813** |  |
| Median (IQR) | 179.2 (118.5 - 222.3) |  | 249.6 (169.7 - 318.0) |  | 376 (253 - 533.0) |  | 362 (246.0 - 510.0) |  | 369.2 (248.0 - 526.8) |  |
| CD4 <200 | 17 | 70.8 | 363 | 34.0 | 4824 | 15.7 | 587 | 16.5 | 5204 | 16.4 |
| CD4 200 – 350 | 5 | 20.8 | 553 | 51.7 | 8856 | 28.3 | 1113 | 31.3 | 9414 | 29.6 |
| CD4 >350 | 2 | 8.3 | 153 | 14.3 | 17040 | 55.5 | 1857 | 52.1 | 17195 | 54.1 |

IQR-interquartile range
†HIV diagnosis is based on point-of-care tests that are not recorded electronically. No HIV evidence therefore includes those who tested negative on point-of-
care tests and therefore have no subsequent electronic record of HIV.

With respect to PCR testing policy periods, 29.1% of pregnancies were during policy period 1 (January 2010 – January 2014), 13.2% in period 2 (August 2014 to November 2015) and 57.7% in period 3 (December 2015 to December 2020).

The proportion of WLWH initiating ART prior to pregnancy increased over time. In 2020, 5.9% (1 103/18 665) of pregnant women with HIV had not initiated ART prior to or during the antenatal period compared to 41.3% (1 568/3 797) in 2010 (Figure 3).

**Figure 3:**
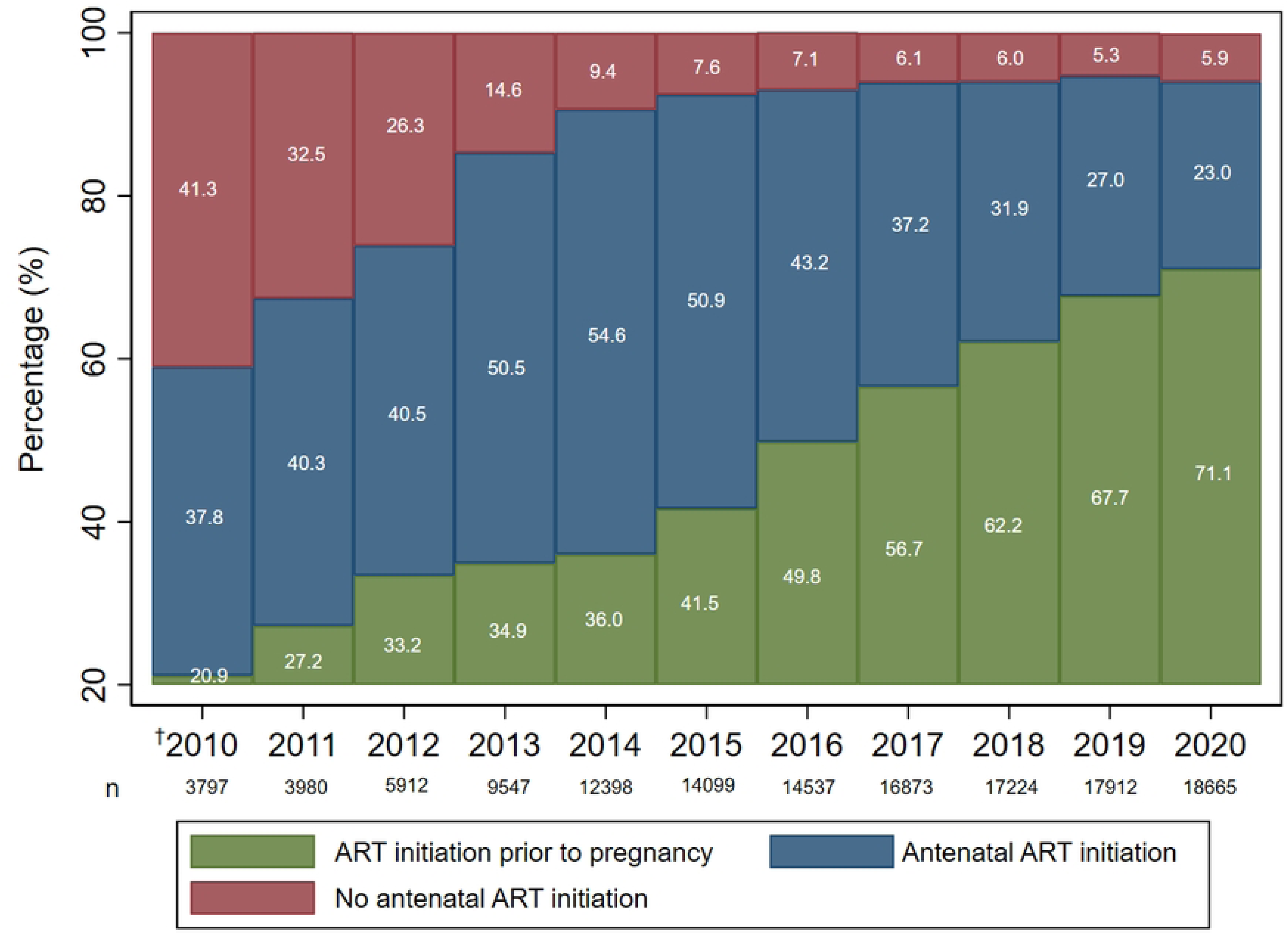
Proportion of maternal antenatal antiretroviral therapy (ART) initiation 2010 – 2020. ^†^**Policy period 1**: Jan – Mar 2010; **policy period 2**: Apr 2010 – Apr 2013; **policy period 3**: May 2013 - Dec 2020

### Vertical transmission of HIV

Of the 842 641 pregnancies, 137 572(16.3%) had a record of child HIV status. Of these, 133 606 (97.1%) children were HIV negative and 3 966 (2.9%) HIV positive by 24 months. Amongst the 3966 children with HIV, median time from birth to HIV diagnosis was 181 days (IQR 33 – 603), and 3 083 (77.7%) were diagnosed with at least one positive HIV PCR test. HIV status of the remaining 883 (22.3%) children with HIV was determined by HIV ELISA testing for children above 18 months of age, or other evidence of HIV where PCR data were not available (for example ART details). Of these 883 children, only 10.2% (90) were diagnosed early (before 7 weeks of age). Among all children with HIV, the percentage of early diagnosis was higher in maternal ART policy period 3 (33.9%) compared to policy periods 1 (29.8%) and 2 (23.7%). This is likely reflective of earlier routine infant PCR testing in maternal ART policy period 3.

Figure 4 shows that while the proportion of children with HIV remained low, the proportion of children exposed with unknown HIV status decreased whilst those testing HIV negative increased. Of the 3 966 children with HIV, 73.2% had evidence of initiating ART.

**Figure 4:**
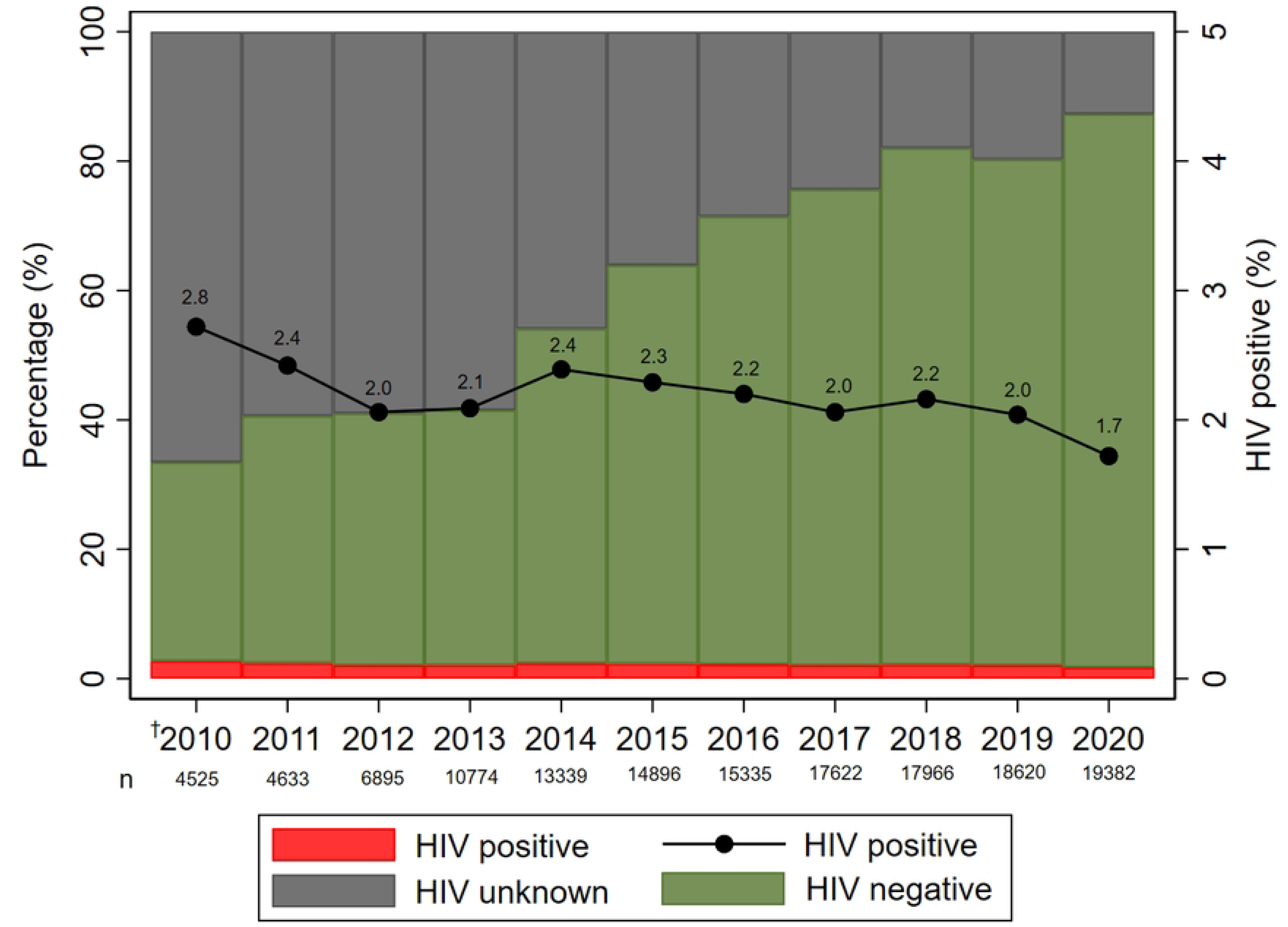
HIV status at 24 months among children HIV exposed 2010 – 2020. ^†^**Policy period 1**: Jan – Mar 2010; **policy period 2**: Apr 2010 – Apr 2013; **policy period 3**: May 2013 - Dec 2020

Overall, 390 (9.8%) of the 3966 children with HIV had apparent negative maternal exposure (electronic record of negative HIV status during pregnancy up to 2 years after delivery) and 517 (13.0%) had unknown maternal exposure. Figures 5a and b summarise maternal and child HIV status over the full study period and more recently (2017 – 2020), respectively. Of all pregnancies, 17.1% (143 987) had known maternal HIV exposure. Of these, 2.1% (95% CI 2.1 – 2.2) had a child diagnosis of HIV, 65.9 % (95% CI 65.7 – 66.2) were HIV negative and 32.0% (31.7 – 32.2%) had no HIV diagnosis (Figure 5a).

**Figure 5a:**
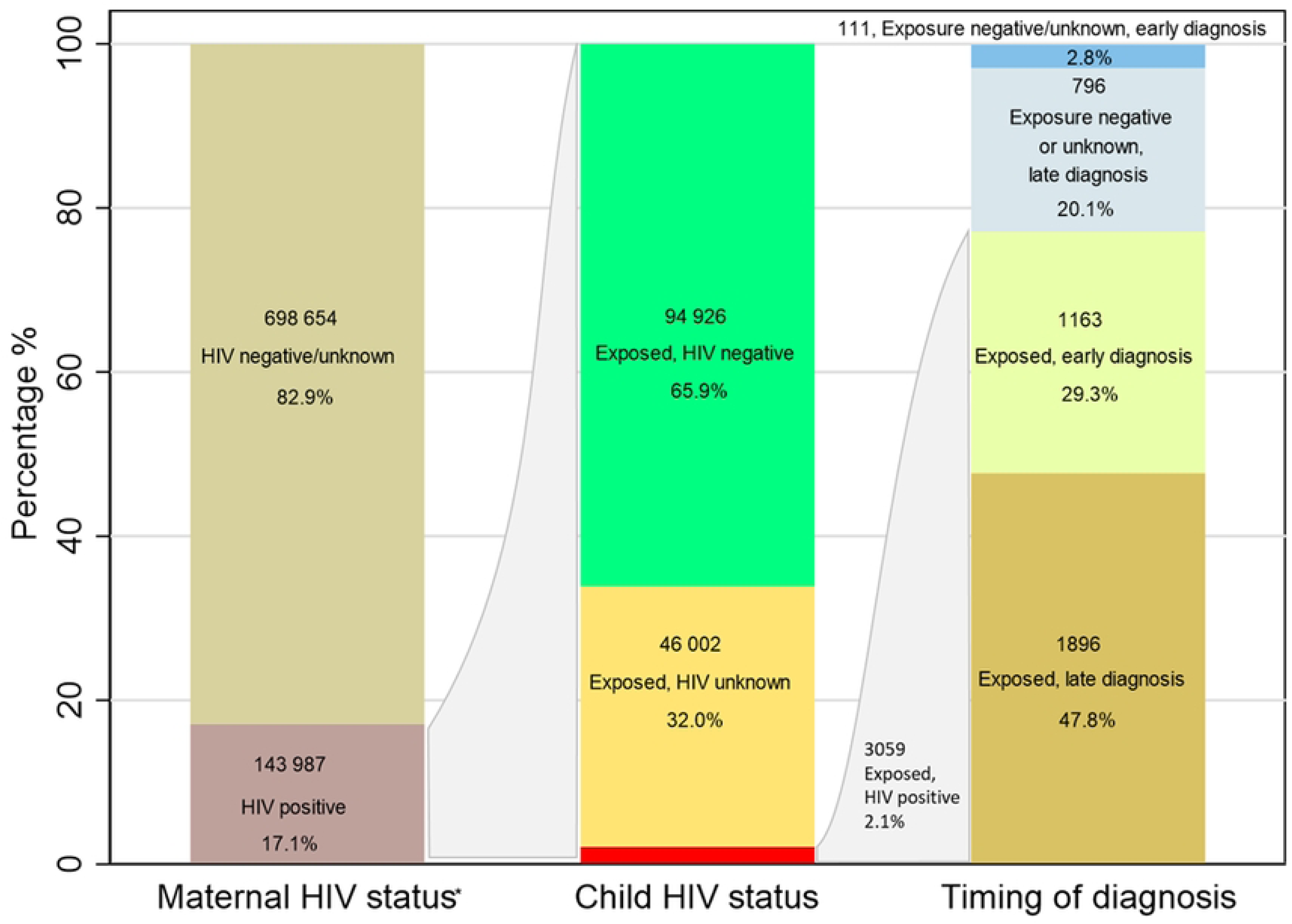
Vertical transmission characteristics (2010 – 2020) *Maternal HIV status is mother’s HIV status during the index pregnancy or within 2 years of delivery

**Figure 5b:**
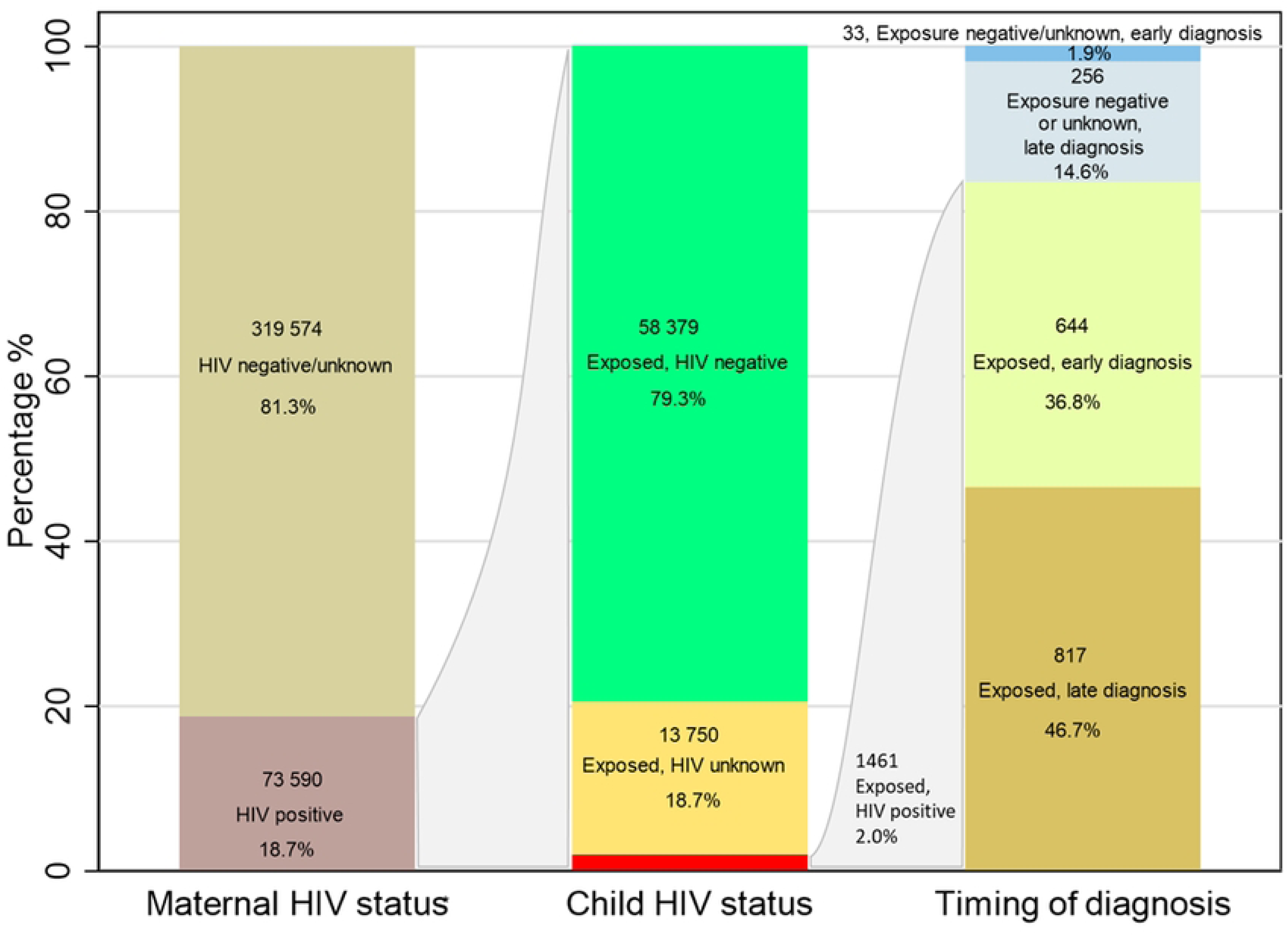
Vertical transmission characteristics (2017 – 2020) *Maternal HIV status is mother’s HIV status during the index pregnancy or within 2 years of delivery

### Factors associated with vertical transmission of HIV

Children born in maternal ART policy period 3 were less likely to be diagnosed with HIV than children born in policy period 2 (aOR 0.66; 95% CI 0.60 – 0.72) (Model 1, Table 2). Across all periods, younger maternal age, lack of maternal antenatal ART, a previous tuberculosis (TB) diagnosis and no record of antenatal visits prior to delivery were associated with vertical transmission of HIV (Table 2). Comparing models 1 and 2, Table 2 shows that the association with maternal ART policy period is mediated by maternal ART use. The impact of suppressive antenatal ART is clearly demonstrated in policy period 3 with those not on ART, 14.44 (95% CI 12.90 – 16.16) times more likely to have vertical transmission of HIV than those on suppressive ART (Model 3).

**Table 2:** Comparison of different models for factors associated with vertical transmission of HIV based on routine data.

|  | Policy periods 1 - 3 |  |  | Policy period 3 |  |
| --- | --- | --- | --- | --- | --- |
| Variable† | OR (95% CI) | Model 1<br>aOR (95% CI) | Model 2<br>aOR (95% CI) | Model 3<br>aOR (95% CI) | Model 4<br>aOR (95% CI) |
| **Maternal ART Policy period** |  |  |  |  |  |
| Policy period 1 | 1.33 (0.98-1.81) | 1.29 (0.95-1.76) | 1.06 (0.75-1.50) |  |  |
| Policy period 2 | 1.00 | 1.00 | 1.00 |  |  |
| Policy period 3 | **0.59** (0.54 -0.64) | **0.66** (0.60-0.72) | **1.12** (1.00-1.24) |  |  |
| **Maternal age category (years)** |  |  |  |  |  |
| 15 - 19 | **1.42** (1.26-1.59) | **1.46** (1.30-1.63) | **1.67** (1.46-1.91) | **1.35** (1.16-1.59) | **1.37** (1.17-1.61) |
| 20-24 | **1.24** (1.15-1.34) | **1.29** (1.20-1.39) | **1.31** (1.20-1.42) | **1.17** (1.07-1.29) | **1.18** (1.07-1.31) |
| 25-39 | 1.00 | 1.00 | 1.00 | 1.00 | 1.00 |
| >39 | 0.82 (0.67-1.00) | 0.80 (0.66-0.98) | **0.78** (0.62-0.98) | 0.86 (0.67-1.10) | 0.86 (0.67-1.10) |
| **Electronic evidence of prior pregnancies** |  |  |  |  |  |
| 0 | 1.00 |  |  |  |  |
| 1 | **0.90** (0.83-0.97) |  |  |  |  |
| 2 | **0.83** (0.75-0.93) |  |  |  |  |
| 3 | 0.90 (0.74-1.08) |  |  |  |  |
| ≥4 | 0.96 (0.70-1.32) |  |  |  |  |
| **Maternal ART** |  |  |  |  |  |
| ART prior to pregnancy | 1.00 |  | 1.00 |  |  |
| Antenatal ART | **1.24** (1.14-1.36) |  | **1.31** (1.20-1.43) |  |  |
| No antenatal ART | **6.43** (5.91-7.00) |  | **4.82** (4.40-5.28) |  |  |
| **No recorded antenatal visits** | **1.72** (1.60-1.84) | **1.57** (1.47-1.69) | **1.36** (1.25-1.47) | **1.13** (1.02-1.24) | **1.13** (1.02-1.24) |
| **Metropolitan district** | **0.75** (0.70-0.80) | **0.85** (0.79-0.91) | 0.94 (0.86-1.02) | 0.99 (0.91-1.09) | 0.99 (0.90-1.08) |
| **History of TB** | **2.16** (2.01-2.32) | **2.14** (1.99-2.29) | **2.18** (2.02-2.35) | **1.85** (1.69-2.02) | **1.79** (1.64-1.96) |
| **Viral Load and ART status ‡** |  |  |  |  |  |
| Suppressed, ART before/during pregnancy | **1.00** |  |  | **1.00** | **1.00** |
| Viraemic, ART before/during pregnancy | **5.17** (4.66-5.74) |  |  | **4.75** (4.27-5.28) | **4.56** (4.10-5.08) |
| VL unavailable, ART before/during pregnancy | **4.10** (3.63-4.64) |  |  | **3.93** (3.47-4.45) | **3.90** (3.44-4.42) |
| VL unavailable, no antenatal ART record | **15.60** (14.01-17.38) |  |  | **14.44** (12.90-16.16) | **14.31** (12.75-16.07) |
| **Last CD4 before booking value (cells/µl) ‡** |  |  |  |  |  |
| CD4 < 200 | 1.00 |  |  |  | 1.00 |
| CD4 200 – 350 | **0.66** (0.52-0.84) |  |  |  | 0.87 (0.68-1.11) |
| CD4 >350 | **0.32** (0.26-0.40) |  |  |  | **0.52** (0.42-0.65) |
| CD4 result not available | **0.54** (0.45-0.65) |  |  |  | **0.65** (0.53-0.79) |

OR - Odds Ratio, aOR - Adjusted Odds Ratio, CI - confidence interval
‡**Outcome variable is infant HIV status at 24 months.** Model 1 includes maternal ART policy period without adjustment for maternal ART, VL and CD4 count which are all impacted by the policy period. Model 2 includes both maternal ART policy period and maternal ART status. Model 3 includes VL status combined with ART status into a composite variable. Model 4 includes combined VL and ART status as well as CD4 status. Patients with VL status but no corroborating ART records were included in Models 1 and 2 but dropped from the models 3 and 4 (effect sizes remain unchanged).
‡Restricted to maternal ART policy period 3

A sensitivity analysis excluding infant deaths during the study period (8712) did not show major impacts on model effects or child HIV trends.

## Discussion

Our findings illustrate temporal trends in vertical transmission of HIV over three maternal ART policy periods in the Western Cape province, with an increasingly greater proportion of WLWH initiating ART prior to pregnancy, and declining vertical transmission of HIV over a 10-year period.

As we used routine data to assemble the study cohort, changes to the cohort over time may be related to variations in availability of routine electronic data. For example, the proportion of pregnancies from non-metropolitan districts was likely lower in earlier years as electronic evidence of pregnancy was limited due to less robust electronic systems in earlier years. Evidence of maternal HIV positive status was stable over time with estimates in earlier years higher than expected and estimates in later years more aligned with antenatal HIV seroprevalence surveys[25,26]. This likely reflects more complete electronic records for WLWH in earlier years compared to those with no evidence of HIV.

Evidence of negative HIV status is low as HIV diagnosis is primarily based on point-of-care HIV tests that are not electronically captured[27]. Amongst those testing positive, electronic records of HIV diagnosis and management after point-of-care diagnosis enable enumeration within the HIV cascade. Since antenatal HIV testing coverage in the Western Cape is high and a recent study reporting digitised point-of-care HIV test results showed the majority of pregnant WLWH testing negative, those with no evidence of HIV likely include a large proportion of women who tested HIV negative in point-of-care tests[27–29]. The increasing proportion of maternal HIV diagnosis before pregnancy supports improved HIV ascertainment. The proportion of women testing HIV positive after pregnancy was lower in later years. This may be due to improved antenatal and pre-pregnancy diagnosis as well as there being less time within the maternal cohort for later pregnancies (cohort bias).

Trends in viral load and CD4 results reflect changing policy, with more viral load testing and fewer CD4 results available in later years illustrating the evolving guidance for more regular viral load testing in pregnancy, and CD4 count testing no longer being indicated for patients stable on ART, as well as earlier initiation of ART following implementation of Option B+ in 2013[30,31]. We found an expected increase in median CD4 count at pregnancy start over time as access to maternal ART increased.

Temporal trends in ART initiation are progressing steadily in that greater proportions of WLWH were initiated on ART prior to or during pregnancy in later years. However, our study showed lower percentages of ART initiation prior to pregnancy in recent years (63.9% in 2019) compared to a smaller study in the Khayelitsha subdistrict (77.9% in 2017)[16]. This difference may be expected given that Khayelitsha has the longest-standing ART programme in the Western Cape, but highlights the need for improved efforts to ensure all service opportunities to test and treat WLWH are utilised appropriately[13]. Of concern, however, is the proportion of women with no evidence of ART in the most recent years pre-COVID (e.g., 5.3% in 2019). This may be due to loss to follow-up for various reasons including migration to a different province as well as incomplete electronic data capturing. These reasons may also apply to children HIV exposed with unknown HIV status, which, although decreasing over time, remains high at 18.7% in recent years (2017 – 2020).

The median age at HIV diagnosis in children of 181 days suggests either late transmission through breastfeeding or delayed diagnosis in many children. Since birth PCR records were not available for all children, exact timing of transmission cannot be discerned, but the lower proportion of late diagnosis in later years suggests improved access to HIV testing in PCR testing policy period 3. A recent study, however, notes that receipt of birth PCR testing from 2015 onwards decreased the likelihood of returning for follow-up testing, which may result in delayed diagnosis[5]. With improved VTP programmes over time, a need for improvement in postnatal VT has been highlighted in several studies[23,24,32]. The overall incidence of VT amongst those with known HIV exposure was 2.1%, comparable to findings in recent South African literature[16,33]. However, a considerable proportion of children with HIV had apparent negative (9.8%) or unknown (13.0%) HIV exposure. These may represent false negative maternal HIV results, missed opportunities for repeat maternal HIV testing, linkage to care and management during pregnancy, or women who may have been diagnosed and managed in other provinces. The decrease in the proportion of children HIV exposed with unknown HIV status and the corresponding increase in children with evidence of negative HIV status suggests temporal trends are understated as undiagnosed child HIV is less likely over time. For example, if a greater proportion of undiagnosed children were HIV positive in earlier years, there may have been a steeper decline in HIV positivity over time than reflected in this analysis.

Of concern is that 26.8% of children with HIV had no evidence of ART initiation. While this may again reflect incomplete electronic records or migration to other provinces, it is important to explore whether there were missed service opportunities for linkage to care, and to explore mechanisms to strengthen follow-up and record these actions electronically.

While our findings clearly demonstrate the impact on VT of evolving guidelines with expanded ART access in pregnancy, the importance of early suppressive ART initiation or intensification was also evident. This supports the urgent need to strengthen access to HIV testing and early antenatal care amongst all women to ensure prompt initiation of ART. In keeping with similar studies, findings from multivariable modelling demonstrate the impact of viraemia (VL≥1000) on vertical transmission, highlighting the urgent need to initiate ART in all eligible women and monitor viral load appropriately[1,14,33]. Although CD4 count, viral load and ART are correlated, the association between CD4 count and vertical transmission persisted after adjustment, with a decreased risk of vertical transmission at higher CD4 counts.

The risk of vertical transmission of HIV decreases with maternal age, with those aged 15 – 19 years most likely to have a child with HIV. This may be because younger age groups are more likely to be diagnosed with HIV during pregnancy, rather than before, following acquisition during recent unprotected sexual encounters[27,34,35]. Furthermore, risk factors for adolescent HIV acquisition and pregnancy may also be risk factors for poor access to health care, including access to HIV testing. Diagnosis and treatment are further delayed if access to antenatal care is delayed, hence exposing the unborn infant to higher viral loads and increasing the risk of vertical transmission[1]. The association between no antenatal care before delivery and vertical transmission is highlighted, supporting findings from similar studies[1,14,33]. The notably lower proportion of pregnancies without antenatal care in policy period 3, and specifically 2019, suggest improved access to antenatal care as well as improved electronic record-keeping during pregnancy in later years. However, targeted interventions for adolescents and young women on regular HIV testing, HIV prevention, including safe sexual practices, and early antenatal care may be of benefit to address the high levels of vertical transmission in younger age groups[2,33,36]. Previous TB was found to be another notable factor associated with vertical transmission. Although timing of TB disease in relation to HIV diagnosis and pregnancy were not evaluated in our study, this association may represent the association of co-morbidities with maternal health through immunosuppression, increased viral load and drug interactions, and as proxies for unmeasured socio-economic associations[37].

## Limitations

Pregnancies from 2018 were evaluated for less than 24 months thus allowing fewer opportunities for ascertainment of HIV diagnosis and management in both mother and child. Since child HIV status was not known for all children who died during the study period, HIV trends may be impacted in either direction. While routine linked individuated information systems provide large, representative datasets for temporal trend and risk factor analysis, these data are prone to administrative errors, incompleteness, varying levels of data quality and contextual factors such as staff training, performance and turnover, disease impacts such as the COVID-19 pandemic, social unrest and natural disasters. Data captured within paper-based systems, such as point-of-care HIV tests, are not integrated into routine electronic data systems. This prevents a clear picture of HIV status amongst those testing negative or testing positive but without records of linkage to HIV care. This study uses ART dispensing as a proxy for ART administration and adherence, however dispensing of ART does not equate to patient adherence. Furthermore, treatment interruption during pregnancy was not evaluated. Even with adjustment for confounding, residual confounding may still impact factors associated with vertical transmission. It should also be noted that these risk factors, described using observational epidemiological methods, are not causal. Since this study was limited to routinely collected data, potential confounding by factors such as socio-economic status, infant feeding methods and co-morbidities (other than previous TB) were not evaluated.

## Conclusions

Our study demonstrates the positive impact of maternal ART policy change and universal test-and-treat to reduce vertical transmission amongst WLWH. Despite the improvements in maternal ART initiation prior to pregnancy and progress towards prevention of vertical transmission, the higher risk of vertical transmission amongst adolescents and young women, those without antenatal care and those with previous TB highlights the potential value of targeted interventions. Our findings emphasise the need for improved methods of support and follow-up amongst pregnant and breastfeeding women with HIV to ensure that all service opportunities to prevent vertical transmission and appropriately manage HIV in both mother and child are utilised optimally.

## Data Availability

The Western Cape Provincial Health Data Centre data used in the study include unconsented, de-identified routine service data housed by the Western Cape Department of Health and Wellness. Release of these data to a public domain would violate the Data Access Policy of the Western Cape Department of Health and Wellness. Ethically approved data requests which may be targeting the same or similar data sources may be sent to or discussed with the Western Cape Provincial Department of Health and Wellness.

## Acknowledgements

The authors gratefully acknowledge the Western Cape Provincial Health Data Centre and the Measurement and Surveillance of HIV Epidemics (MeSH) consortium. We also acknowledge the funders of this work, Bill & Melinda Gates Foundation https://www.gatesfoundation.org/ (BR, AB and NJ were recipients of funding from grant OPP1191327).

## Notes

### Competing Interest Statement

The authors have declared no competing interest.

### Funding Statement

Yes

### Author Declarations

The study was approved by the University of Cape Town Human Research Ethics Committee (HREC 083/2021) and the Western Cape Provincial Health Research Committee.

